# Change in body weight of older adults before and during the COVID-19 pandemic: Longitudinal results from the Berlin Aging Study II

**DOI:** 10.1101/2023.09.08.23295246

**Authors:** Valentin Max Vetter, Johanna Drewelies, Sandra Düzel, Jan Homann, Lil Meyer-Arndt, Julian Braun, Anne Pohrt, Friederike Kendel, Gert G. Wagner, Andreas Thiel, Lars Bertram, Vera Regitz-Zagrosek, Denis Gerstorf, Ilja Demuth

**Affiliations:** Charité – Universitätsmedizin Berlin, corporate member of Freie Universität Berlin and Humboldt-Universität zu Berlin, Department of Endocrinology and Metabolic Diseases (including Division of Lipid Metabolism), Biology of Aging working group, Augustenburger Platz 1, 13353 Berlin, Germany; Lise Meitner Group for Environmental Neuroscience, Max Planck Institute for Human Development, Berlin, Germany; Department of Psychology, Humboldt University Berlin, Berlin, Germany; Max-Planck Institut für Bildungsforschung; Friede Springer Cardiovascular Prevention Center; Charité – Universitätsmedizin Berlin (CBF), Berlin, Germany; Institute of Epidemiology and Social Medicine, University of Münster, Münster, Germany; Charité - Universitätsmedizin Berlin, corporate member of Freie Universität Berlin, Humboldt-Universität zu Berlin, and Berlin Institute of Health, Regenerative Immunology and Aging, BIH Center for Regenerative Therapies, 13353 Berlin, Germany; Charité – Universitätsmedizin Berlin, corporate member of Freie Universität Berlin, Humboldt-Universität zu Berlin, and Berlin Institute of Health, NeuroCure Clinical Research Center, 10117 Berlin, Germany; Charité – Universitätsmedizin Berlin, corporate member of Freie Universität Berlin, Humboldt-Universität zu Berlin, and Berlin Institute of Health, Department of Neurology with Experimental Neurology, 10117 Berlin, Germany; Department of Medical Biometrics, Charité - Universitätsmedizin Berlin, corporate member of Freie Universität Berlin, Humboldt-Universität zu Berlin, and Berlin Institute of Health, Berlin, Germany; Gender in Medicine, Charité - Universitätsmedizin Berlin Corporate Member of Freie Universität Berlin and Humboldt-Universität zu Berlin, Berlin, Germany; German Socio-Economic Panel Study (SOEP), German Institute for Economic Research (DIW), Berlin, Germany; Center for Lifespan Psychology, Max Planck Institute for Human Development, Berlin, Germany; Lübeck Interdisciplinary Platform for Genome Analytics (LIGA), University of Lübeck, Lübeck, Germany; Institute for Gender in Medicine, Center for Cardiovascular Research, Charité – Universitätsmedizin Berlin, Corporate Member of Freie Universität Berlin, Humboldt— Universität zu Berlin and Berlin Institute of Health, Berlin, Germany; Department of Cardiology, University Hospital Zürich, University of Zürich, Zürich, Switzerland

## Abstract

**Background:** Change in body weight during the COVID-19 pandemic as an unintended side effect of lockdown measures has been predominantly reported for younger and middle-aged adults. However, information on older adults for which weight loss is known to result in adverse outcomes, is scarce.

**Aims:** Describe body weight change in older adults before, during, and after the COVID-19 lockdown measures and explore putative associated factors with a focus on the period that includes the first six months of the COVID-19 containment measures.

**Methods:** In this study, we analyzed the longitudinal weight change of 472 participants of the Berlin Aging Study II (mean age of 67.5 years at baseline, average follow-up time 10 years). Additionally, differences between subgroups characterized by socio-economic, cognitive, and psychosocial variables as well as morbidity burden, biological age markers (epigenetic clocks, telomere length), and frailty were compared.

**Results:** On average, women and men lost 0.87% (n=227) and 0.5% (n=245) of their body weight per year in the study period covering the first six months of the COVID-19 pandemic. Weight loss among men was particularly pronounced among groups characterized by change in physical activity due to COVID-19 lockdown, low positive affect, premature epigenetic age (7-CpG clock), diagnosed metabolic syndrome, and a more masculine gender score (all variables: p<0.05, n=245).

**Conclusions:** Older participants lost weight with a 2.5-times (women) and 2-times (men) higher rate than what is expected in this age.

## Introduction

The outbreak of the novel severe acute respiratory syndrome coronavirus 2 (SARS-CoV-2) was declared to be a public health emergency of international concern on January 30^th^, 2020, by the World Health Organization (WHO) [1]. To contain its spread, non-pharmaceutical interventions (NPIs) were implemented by governments across the world [1-3] as numbers of cases and deaths were rising. They included, among others, closing of schools, social distancing, and lockdowns on the population level [4]. In Berlin, Germany, the first SARS-CoV 2 positive patient was registered on March 1^st^, 2020. Gradually increasing restrictions were subsequently imposed in Berlin and culminated in the first full lockdown in mid-March and April 2020 [5, 6]. A second lockdown was in effect between November 2020 and April 2021. However, the degree of restrictions varied within and across the lockdown periods and different rules applied to persons with a negative Sars-CoV-2 test.

While lockdowns proved, at least for the most part, to be effective in keeping the number of Sars-Cov-2 infections low [3, 4, 7], other effects of NPIs, both beneficial and adverse, were reported. For example, while unfavorable nutritional habits (e.g., an increase in consumption of comfort food and alcohol) seemed to be predominant, beneficial dietary changes (e.g., increased consumption of fresh produce and home cooking) were also reported (reviewed in [8] and [9]). Body weight was one important anthropometric measure that was potentially impacted by these changes in dietary behavior, but potentially also by psychosocial stress, mobility restrictions, individual predispositions, and other factors associated with social restrictions or COVID-19 itself [8, 10-15]. To quantify this effect, several studies analyzing body weight during the time of the COVID-19 pandemic and associated lockdown measures have been published (reviewed in [9, 14-16]). However, almost all of these studies (except for [10, 17]) relied on cross-sectional data only. Furthermore, only two studies examined older adults, here defined as individuals with a mean age above 65 years [11, 17]. This type of data are needed because earlier reports have demonstrated both an increase of malnutrition during lockdown measures among older adults [17] and that low body weight [18] and especially weight loss [18, 19] are associated with higher mortality in this vulnerable group (obesity paradox [18]).

In this study, we aimed to close this knowledge gap by describing the longitudinal change of body weight in participants of the Berlin Aging Study II (BASE-II [20]) before, during, and after the time COVID-19 lockdown measures were implemented. In a second step, we explore differences in body weight change between subgroups characterized by variables known or hypothesized to be associated with healthy aging or body weight.

## Methods

### Study Population

The Berlin Aging Study II (BASE-II) is a multicenter interdisciplinary study with the goal to identify health-promoting factors in old age [20]. Baseline examination (T0) of 1,671 older participants in the medical part of the study (mean age: 68.8 years, sd: 3.7 years, 51.6% women) was conducted between 2009 and 2014. Between 3.9 and 10.4 years later (mean 7.4 years, sd = 1.5 years) a follow-up examination of 1,083 participants (+ 17 additional participants) as part of the GendAge study (T1) [21] was conducted. This examination (T1) took place between June 22^nd^ 2018 and March 10^th^ 2020, thereby was completed shortly before the start of local COVID-19-restrictions. Six months into the COVID-19 pandemic an online survey was sent out to the participants. At the time the online survey was conducted, none of the participants reported to have had a COVID-19 since the beginning of the pandemic. In the final dataset, 472 participants provided information on their body weight at T0, T1 and online survey (Figure 1B).

**Figure 1:**
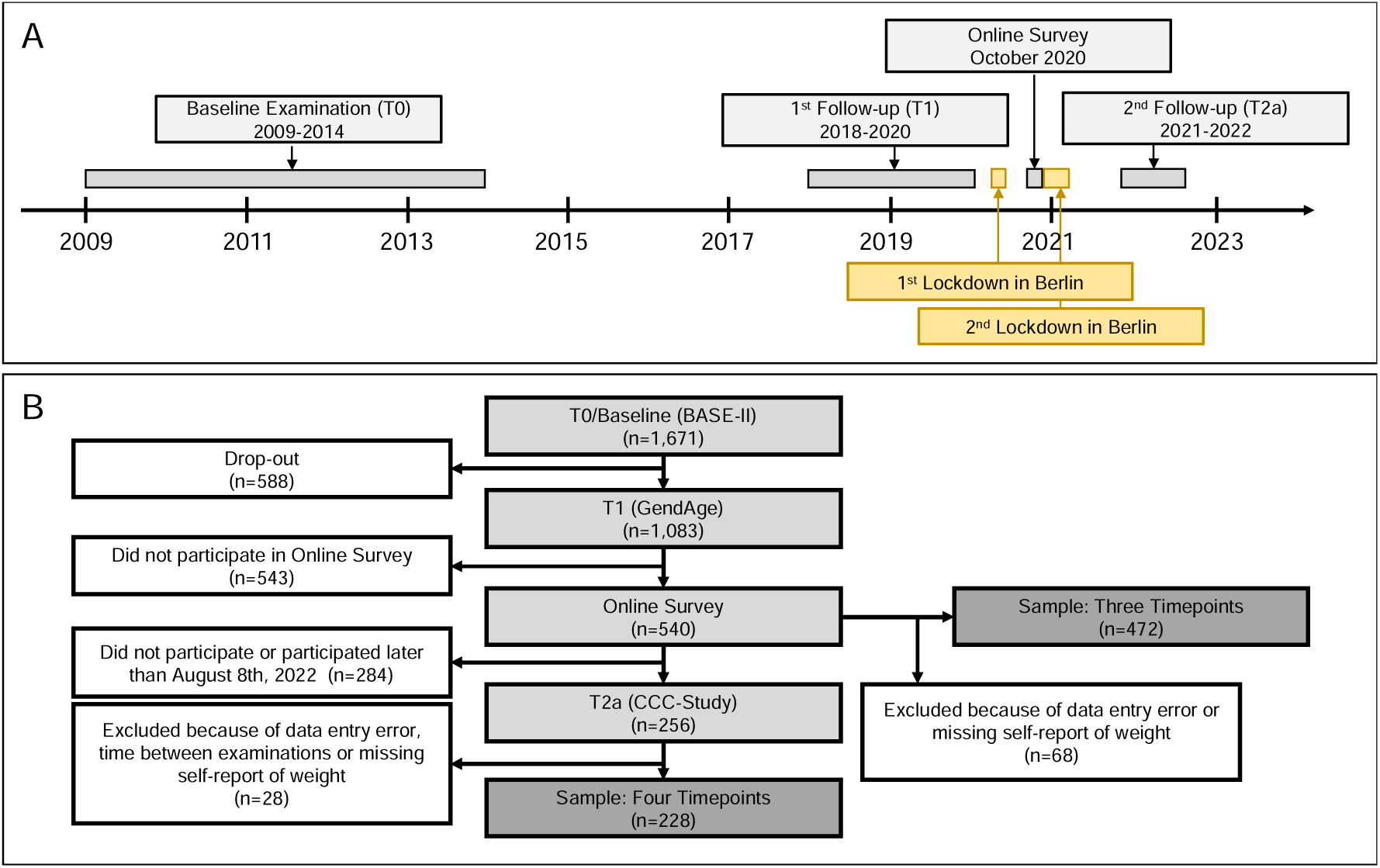
A: Timeline visualizing baseline, first follow-up examination, Online Survey and second follow-up examination of the BASE-II cohort. B: Flowchart illustrating how the final sample including three and four timepoints was defined.

A second medical follow-up examination (T2) began in February 2021 and was completed in July 2023 as part of the Charité Corona Cross study. Because we focused on COVID-19- pandemic lockdown associated changes in this study, we included only participants who were examined before August 8^th^, 2022 (T2a) in our analyses. However, this choice was arbitrary as no meaningful cut-off is available. A sample of 228 participants was available that provides information on all four time-points (T0, T1, online survey, and T2a, see Figure 1B). Potential bias due to non-random loss to follow-up was examined (Supplementary Table 1). If any, only very limited selection bias is observable in the variables examined here. This manuscript was created in accordance with the STROBE guidelines [22, 23].

### Body weight

Participants were asked to report their body weight as part of a questionnaire (in kilogram with one or no decimal places). Additionally, body weight was measured at T0 and T1 (after the self-reported weight has been recorded) with the electronic measuring station seca 763 (SECA, Germany) in kilogram with two decimal places. To improve comparability between all four timepoints, the self-reported weight was used for all subsequent analyses. The participant’s individual weight change between two consecutive timepoints was calculated as annual percentage change making use of an exponential growth function known as compound annual growth rate (CAGR):

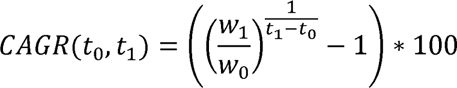

where w_0_ is the initial weight (in kg) at timepoint t_0_ which is followed by the consecutive measurement of body weight w_1_ at timepoint t_1_. The time difference between body weight assessments t_1_ – t_0_ was calculated as difference between age in years at time of assessment.

### Additional Variables

All additional variables, except self-reported change in physical activity (PA) due to COVID- 19, were assessed at the first follow-up examination (T1) that was completed just before the pandemic started. A more in-depth description on how these variables were assessed can be found in the Supplementary Methods and Supplementary Table 2.

### General Characteristics

Biological sex was assessed dichotomously as "man" or "women". To separate the effects of socially constructed gender from sex a "gender score" was recorded. This score aggregates information on central gender dimensions. Lower values indicate more traditionally male characteristics (for more details see [24]). Cohabitation was derived from several questions about the participant’s household and living situation (further information in Supplementary Methods). BMI at T1 was calculated as measured weight (in kg) divided by squared height (in m). Education was measured in number of years (7 to 18 years). Income was assessed in eighteen categories as part of the self-administered questionnaire.

### Physical Activity (PA)

Participants were asked to rate themselves as “active” or “rarely/never active” (reported as “not active” of the variable “Physically Active” in this study) as part of the Rapid Assessment of Physical Activity (RAPA) [25] questionnaire. Additionally, PA was measured with the wGT3X-BT activity monitor (ActiGraph LLC, USA) and reported as Vector Magnitude (VM) count [26]. Participants were asked to report change in their PA that specifically needs to be attributed to the CVOID-19 pandemic as part of the online survey six months into the pandemic.

### Cognitive performance, Hand Grip Strength and Frailty

Cognitive performance, specifically perceptual speed, was assessed by the Digit Symbol Substitution Test (DSST) [27]. Muscle strength was assessed as hand grip strength, and pre-sepcified cut-off values were used to define impairment [28, 29]. Frailty indices were calculated following the approach by Fried and colleagues [29] as well as the deficit accumulation approach implemented in the SPRINT study [30].

### Cardiovascular Risk and Chronic Disease

Ten-year risk for cardiovascular disease was calculated with the SCORE2 and SCORE2-OP instrument [31, 32] (hereinafter referred to as SCORE2). Type 2 Diabetes Mellitus (T2D) was diagnosed based on the criteria defined by the American Diabetes Association (ADA) guidelines [33]. Metabolic syndrome was diagnosed according to the American Heart Association/ International Diabetes Federation/ National Heart, Lung, and Blood Institute criteria 2009 [34]. Kidney function was determined by estimating the glomerular filtration rate (eGFR) based on cystatin C using the CAPA equation [35]. To get a more general estimation of the participants morbidity, an adapted version [36] of Charlson’s morbidity index [37] was calculated.

### Biomarkers of Aging

The 7-CpG clock [38] and GrimAge clock [39] DNA methylation age (DNAmA) were measured in EDTA blood samples collected at the first follow-up. DNAmA acceleration (DNAmAA) was calculated as leukocyte-adjusted residuals of a linear regression of DNAmA on chronological age (for details please [40]). Telomere length was estimated with a recently published algorithm from epigenome-wide methylation data [41].

### Psychosocial Resources

Affect was assessed by the Positive and Negative Affect Schedule (PANAS-X [42]) [43]. The UCLA Loneliness Scale [44] was used to assess loneliness [45]. A 3-item subscale of the short version of the Big Five Inventory (BFI) [46] was used to assess conscientiousness [47]. Perception of stress was assessed by eight items of the Perceived Stress Scale (PSS) [48]. Participants answered the questions on a scale from 1 (“never”) to 5 (“very often”).

### Missing Values

In the final dataset, 5.2% of data was missing and subsequently imputed with R’s mice package [49] (20 imputed datasets, 5 iterations). Convergence of the imputation algorithm as well as distribution and proportions of imputed variables were inspected visually. As part of a sensitivity analysis, we re-ran our models using the original (not-imputed) dataset which revealed a highly comparable pattern of results (Supplementary Figure 5 and 6).

### Statistical Analyses

All statistical analyses were conducted in R, version 4.2.1 [50] and figures were drawn with R’s “ggplot2” package [51]. Mean difference between groups as well as 95% confidence intervals and p-values for two-sided two-sample t-tests were calculated. Continuous variables were dichotomized to allow a more easy and intuitive comparison.

## Results

### Participants

In this study we analyzed 472 BASE-II participants that provided information on their body weight at three timepoints. After the baseline examination (2009-2014, T0), the first follow-up examination (T1) was conducted on average 7.4 years later and was completed just before the beginning of the pandemic. The third data assessment was conduct as an online survey six months into the pandemic. To examine how the weight of the participants developed subsequently, we analyze a subsample of 228 participants who were followed-up a fourth time on average about 1 year later (T2a).

Mean age at T0, T1, online survey, and T2a were 67.5, 74.8, 76.3, and 77.4 years. About half of the study sample are women (48%). Follow-up time between T0 and T1, T1 and online survey, and online survey and T2a were 7.3, 1.5, and 1.1 years. Self-reported and measured weight correlated highly at T0 (r=0.98) and at T1 (r=0.99, Figure 2 B and C), suggesting a high degree of comparability between self-reported and objectively measured weight.

**Figure 2:**
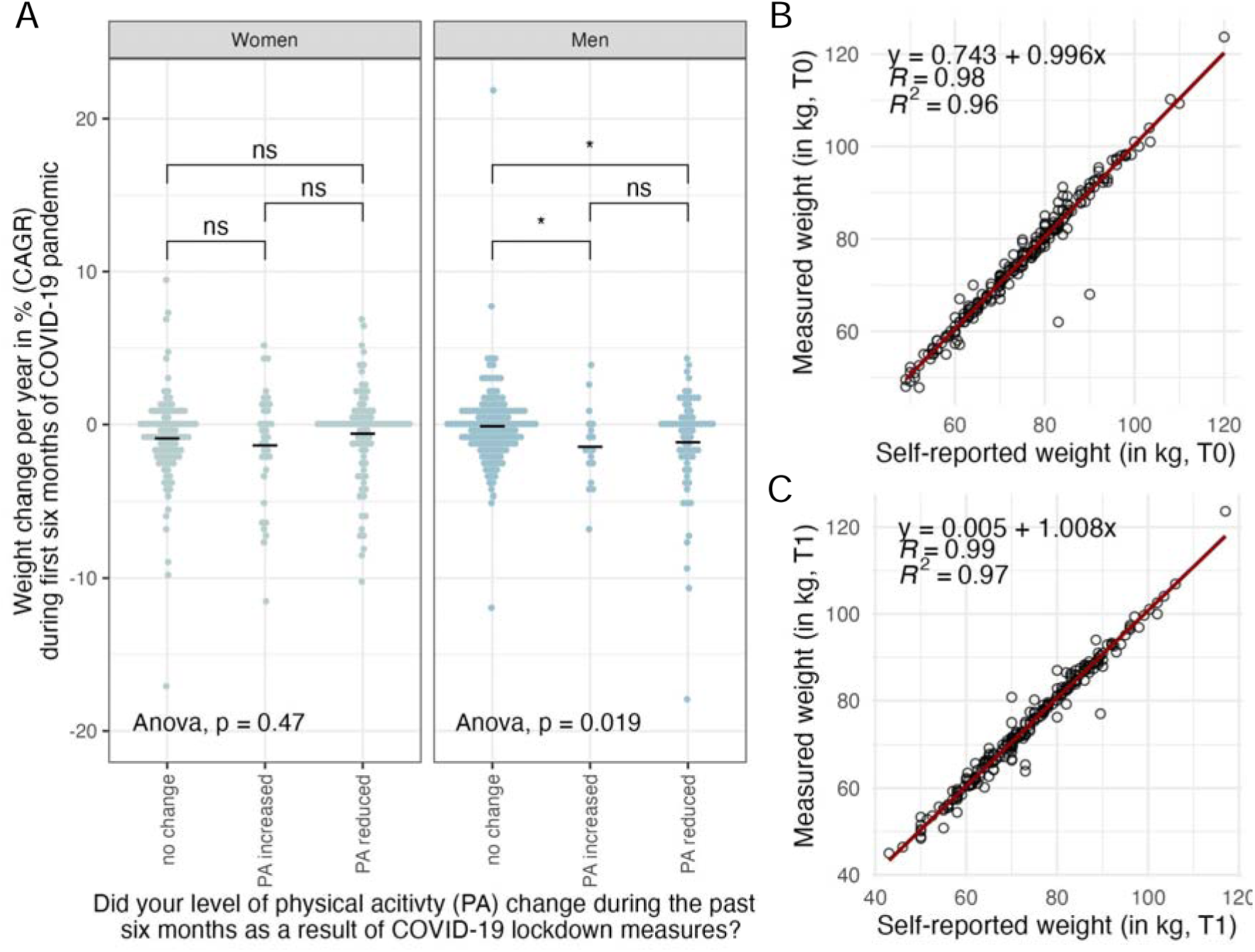
A: Boxplot displaying the difference in weight change stratified by self-reported change in physical activity during the first six months of COVID-19 lockdown. B and C: Scatterplots illustrating the high correlation between measured and self-reported weight during baseline (T0) and first follow-up (T1) examination. Note: kg: kilogram; CAGR: compound annual growth rate, T0: baseline examination, T1: first follow-up examination; Onl. Surv.: Online Survey; T2: second follow-up examination.

### Change in body weight

Mean change of body weight per year in percent (compound annual growth rate, CAGR) between T0 and T1 was -0.17% for men and -0.04% for women (Supplementary Table 2). Between T1 and the online survey (mean follow-up time = 1.5 years, SD = 0.47 years), which is the period that covers the first six months of the COVID-19 pandemic in Berlin as well as associated lockdown measures and restrictions, men lost on average 0.5% and women lost on average 0.87% of their body weight per year. In the period following the online survey in which lockdown measures were generally less strict, men gained 0.93% of body weight while women continued to lose on average 0.02% per year (Table 1, Figure 3). Descriptive statistics of the sample of participants who provided data at only three time points (n=472) is provided in Supplementary Table 2.

**Figure 3:**
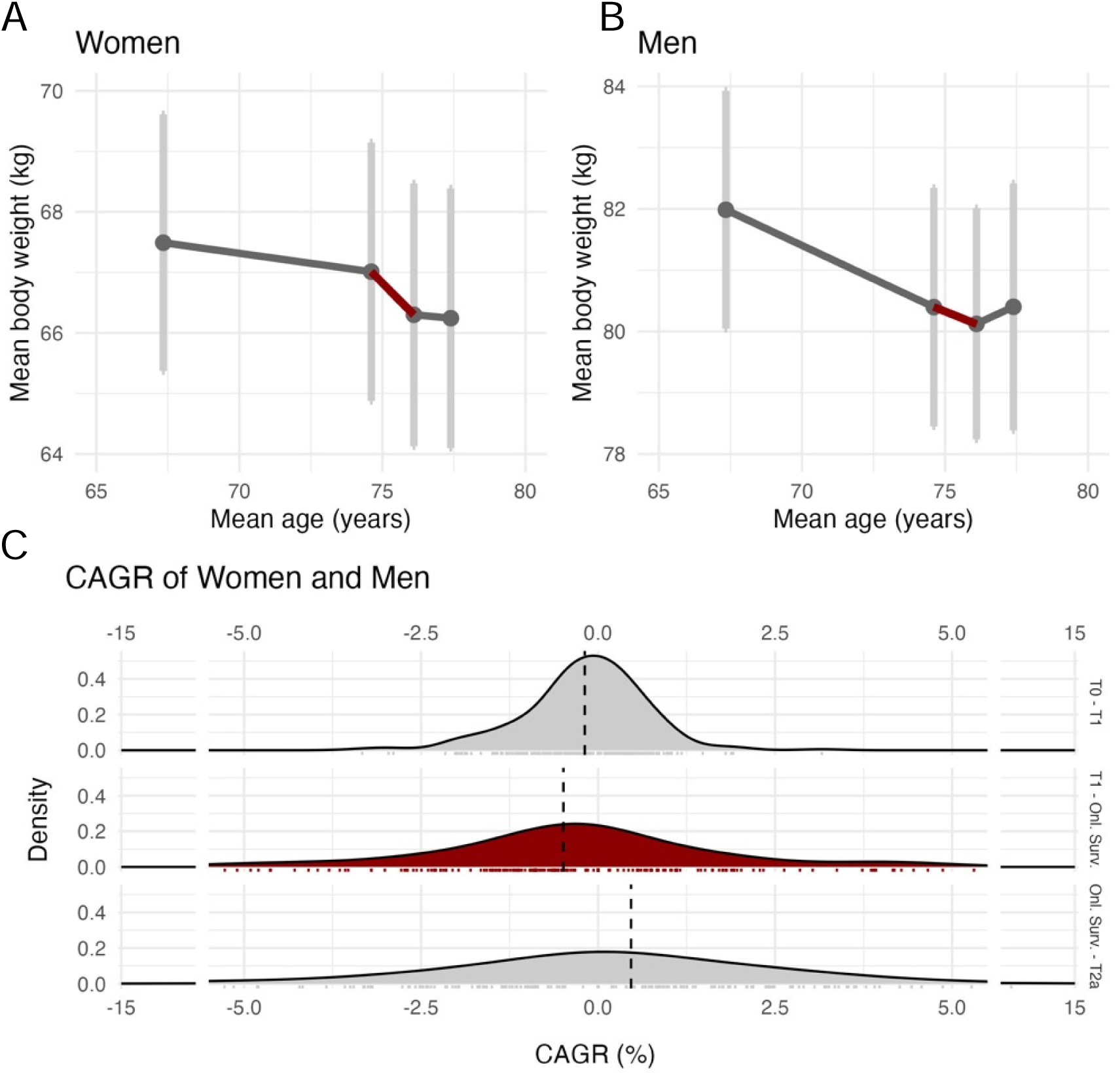
Mean and 95% CI of BASE-II participants body weight at T0, T1, online survey, and T2a (n=227) stratified for subgroups of women (A) and men (B). Density curves of CAGR (Change in body weight per year in percent) between T0/T1, T1/ Online Survey, and Online Survey/T2 for women and men (C). Note: kg: kilogram; CAGR: compound annual growth rate, T0: baseline examination, T1: first follow-up examination; Onl. Surv.: Online Survey; T2: second follow-up examination.

**Table 1:** Descriptive statistics of age and body weight of the subsample of BASE-II participants that provided information at all four timepoints (n=228).

| Timepoint | Variable | Women (n=111) |  |  |  | Men (n=117) |  |  |  | p-value |
| --- | --- | --- | --- | --- | --- | --- | --- | --- | --- | --- |
|  |  | mean | sd | min | max | mean | sd | min | max |  |
| T0 | Age (years) | 67.01 | 2.71 | 61.30 | 73.30 | 67.64 | 3.39 | 60.40 | 77.30 | 0.122 |
|  | Body weight (kg) | 67.49 | 11.27 | 49.00 | 103.30 | 81.99 | 10.60 | 60.50 | 120.00 | 0.000 |
| T1 | Age (years) | 74.64 | 3.08 | 67.10 | 82.70 | 74.58 | 3.80 | 65.80 | 85.90 | 0.895 |
|  | Body weight (kg) | 67.01 | 11.35 | 43.00 | 106.00 | 80.40 | 10.62 | 60.00 | 117.00 | 0.000 |
| Online Survey | Age (years) | 76.10 | 3.05 | 68.60 | 83.60 | 76.10 | 3.79 | 68.20 | 87.70 | 0.994 |
|  | Body weight (kg) | 66.30 | 11.54 | 45.00 | 100.00 | 80.12 | 10.31 | 60.00 | 115.00 | 0.000 |
| T2a | Age (years) | 77.40 | 2.98 | 70.20 | 85.20 | 77.37 | 3.59 | 69.80 | 88.80 | 0.952 |
|  | Body weight (kg) | 66.24 | 11.39 | 43.00 | 103.00 | 80.40 | 11.01 | 61.00 | 130.00 | 0.000 |
| T0 to T1 | Body weight change (kg) | -0.48 | 3.59 | -10.00 | 6.50 | -1.59 | 5.32 | -21.50 | 15.50 | 0.068 |
|  | Body weight change per year (kg/year) | -0.07 | 0.51 | -1.70 | 1.20 | -0.21 | 0.75 | -3.10 | 2.50 | 0.106 |
|  | Body weight change CAGR (%) | -0.11 | 0.77 | -2.90 | 1.90 | -0.26 | 0.91 | -3.30 | 3.20 | 0.170 |
| T1 to Online Survey | Body weight change (kg) | -0.71 | 2.79 | -11.00 | 7.00 | -0.27 | 3.52 | -19.00 | 13.00 | 0.300 |
|  | Body weight change per year (kg/year) | -0.48 | 2.06 | -6.80 | 6.60 | -0.29 | 2.65 | -16.10 | 12.90 | 0.563 |
|  | Body weight change CAGR (%) | -0.74 | 3.08 | -10.20 | 9.40 | -0.25 | 3.42 | -17.70 | 21.80 | 0.259 |
| Online Survey to T2a | Weight change (kg) | -0.06 | 2.74 | -14.00 | 10.00 | 0.28 | 3.76 | -13.00 | 20.00 | 0.446 |
|  | Weight change per year (kg/year) | -0.05 | 2.15 | -10.30 | 7.80 | 0.66 | 4.88 | -8.20 | 43.20 | 0.159 |
|  | Weight change CAGR (%) | -0.02 | 3.10 | -13.80 | 12.10 | 0.93 | 7.37 | -8.60 | 72.10 | 0.212 |

Only very few participants switched weight quartiles between T1, Online Survey, and T2a. This indicates that the weight changes between timepoints were not the result of extreme changes of only a few individuals but result from the change of the whole study population (Supplementary Figure 1).

Cross-sectionally, age was not correlated with body weight in men (r<0.05) and only modestly correlated in women (r= -0.2, Supplementary Figure 2 B). Individual weight trajectories are visualized in Supplementary Figure 2 A.

### Comparison of change in body weight between specific subgroups

To further explore changes in body weight, in a first step we analyzed differences between participants who reported to have changed their physical activity levels due to COVID-19 lockdown measures assessed in an online survey six months into the COVID-19 epidemic and those who reported no change. While no statistically significant weight change differences were found in women (ANOVA, p=0.47), men who reported to have increased (mean= -1.5%, n=18) and reduced PA (mean= -1.2%, n=69) differed statistically significantly in their weight change (CAGR) from men who reported to not have changed their PA (mean=-0.1%, n=157) due to COVID-19 lockdown measures (Figure 2 A).

In a second step we aimed to describe differences in body weight change between subgroups that were defined by variables assessed at the first follow-up examination which was completed just before the first COVID-19 lockdown measures were implemented (T1). Descriptive statistics of analyzed variables can be found in Supplementary Table 1.

In women no statistically significant difference between weight change was found when comparing subgroups. Male participants with lower values in positive affect, premature biological aging (7-CpG epigenetic clock), diagnosed metabolic syndrome, and a lower gender score (more masculine) lost significantly more weight than participants in the comparison group (Figure 4).

**Figure 4:**
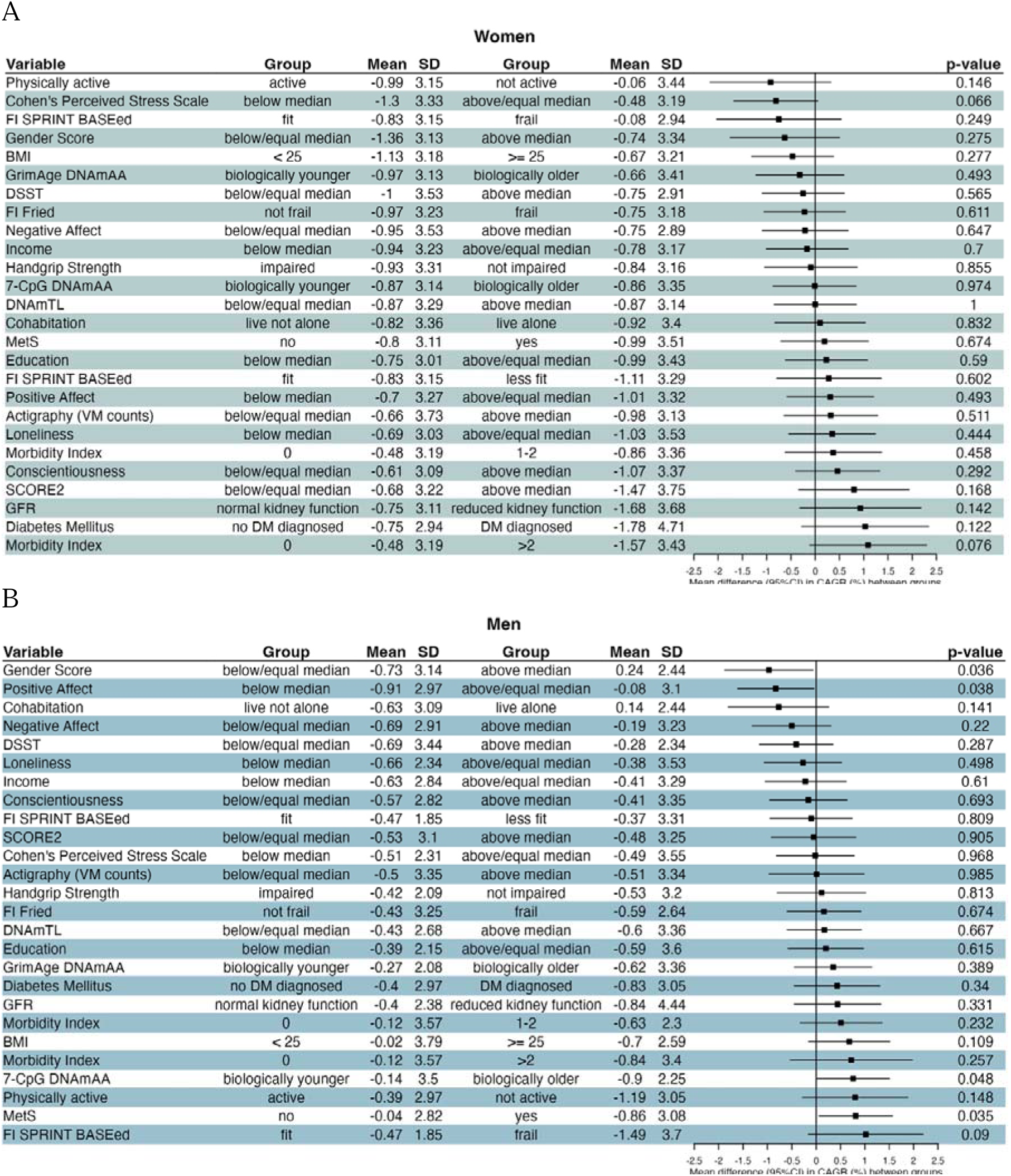
Forest plot of mean difference and 95% CI between CAGR (%) between groups defined by general characteristics, physical activity, geriatric assessments, chronic disease, biomarkers of aging, and psychosocial parameters. Note: DSST: Digit Symbol Substitution Test; DNAmAA: DNA methylation age acceleration; DNAmTL: DNA methylation telomere length; VM: Vector Magnitude; GFR: glomerular filtration rate; BMI: Body Mass Index; MetS: Metabolic Syndrome; FI: Frailty Index; DNAmAA: DNA methylation age acceleration; DNAmTL: DNA methylation telomere length; CAGR: compound annual growth rate.

The combined analysis of women and men is shown in Supplementary Figures 3 and 4. Additionally, Pearson’s correlations coefficients between weight change (between T1 and online survey) and continuously measured variables of interest at T1 are shown in Supplementary Figure 7.

## Discussion

In this study, we assessed the change in body weight before, during and after COVID-19 lockdown measures in an otherwise healthy elderly (age >65years) cohort from Berlin, Germany. Weight loss in the period covering the first six months of the COVID-19 pandemic was 2.5-times (women) and 2-times (men) higher than expected in this age group according to a large dataset of participants from Austria (n=185,192 [52]). Interestingly, after the initial six months of the pandemic, the women’s weight was almost stable (on average -0.02% of body weight per year) and men even gained on average 0.93% of body weight per year eventually reaching their average body weight that was assessed at the examination prior to the pandemic (Table 1). A lower than expected body weight is of concern because it was shown to be associated with increased mortality [18]. Furthermore, unintended weight loss is part of the well-established Fried frailty phenotype [29].

Our findings contrast with the only other study examining a healthy study sample with a mean age of more than 65 years. DiSanto and colleagues who found in a survey conducted with 128 participants between 60 and 87 years that 35% reported weight gain and 11% reported weight loss during the time COVID-19 containment measures were in place[11]. One recent systematic review on the change of body weight and BMI during the first COVID-19 lockdown period identified 36 eligible studies [14]. The authors of the systematic review suggested that the focus on younger people in the literature might be the result of the data collection and recruitment methods which were predominantly online and therefore potentially were less accessible for older adults [15]. This illustrates that, although this topic is of high interest, data on the group of older adults is scarce.

In a second step, we compared change in body weight between different subgroups in our sample. Men who reported to have changed their PA (either increased or reduced PA) due to COVID-19 lockdown measures had lost significantly more body weight compared to those who did not change their PA levels. Furthermore, statistically significant differences in body weight change were found for men between groups characterized by positive affect, epigenetic age (7-CpG clock), metabolic syndrome and socially constructed gender (gender score). We note that these findings result from explorative analyses, and therefore cannot be used for prediction or causal inferences without further independent validation.

There are several limitations to this study. First, as with every longitudinal study, evaluation of potential bias due to loss to follow-up poses a challenge. However, a comparison of the subsample of participants providing information at all three timepoints (n=472) with the complete sample at baseline shows no large deviation (Supplementary Table 1). Second, although body weight of participants was measured with an electronic scale at T0 and T1, only self-reported body weight was available at all four timepoints. However, we are confident that this has only limited impact on the results of our study as the correlation between objectively measured and self-reported weight (which was recorded before body weight was measured) at T0 and T1 was very high. At the same time, we cannot rule out context effects. At the study site, participants may have reported more accurate weights because they expected a subsequent objective assessment, whereas it was clear to them that the online questionnaire would not involve any objective verification. A third limitation to this study is the above-average health, educational level and income of this particular dataset as described before [20] limiting the generalizability of the results. Furthermore, participants of BASE-II might have access to coping strategies that are not available to other parts of the population, resulting potentially in underestimating the adverse effects observed in this study. Fourth, because of the explorative nature of our analyses, we did not adjust for multiple testing. Therefore, an independent validation of our results is needed before final conclusion can be drawn.

Strengths of this study include the well characterized large cohort of older adults. The high quality of assessed data at four examinations of which the second just ended before the first COVID-19 cases were reported in Berlin allows a detailed description of weight change. Additionally, the wide range of variables that is available in the BASE-II allows a comprehensive explorative analysis of different aspects and individual levels of functionality.

## Conclusion

Change in body weight in the study period covering the first 6 months of the COVID-19 pandemic were about 2.5- (women) and 2-times (men) higher than what would be expected in this age group. Body weight change in men differed statistically significantly between subgroups characterized by change of PA due to COVID-19 lockdown, positive affect, biological age (7-CpG clock), metabolic syndrome, and gender score. Although these descriptive results need to be validated independently, they point to interesting candidate markers for future studies.

## Ethics approval and consent to participate

All participants gave written informed consent. All medical assessments were conducted in accordance with the Declaration of Helsinki and approved by the Ethics Committee of the Charité – UniversitaCtsmedizin Berlin (approval numbers EA2/029/09, EA2/144/16, and EA2/224/21) and were registered in the German Clinical Trials Registry as DRKS00009277and DRKS00016157 (BASE-II and GendAge).

## Consent for publication

Not applicable.

### Availability of data and materials

Due to concerns for participant privacy, data are available only upon reasonable request. Please contact Ludmila Müller, scientific coordinator, at, for additional information.

## Author contributions

Conceived and designed the study: V.M.V and I.D. Contributed study-specific data: V.M.V., J.D, S.D., J.H., J.B. L.M.-A., A.P., F.K., G.G.W., A.T., L.B., V.R.-Z., D.G. and I.D. Analyzed the data: V.M.V. Wrote the manuscript: V.M.V and I.D. All authors revised and approved the manuscript.

## Data Availability

Due to concerns for participant privacy, data are available only upon reasonable request. Please contact Ludmila Mueller, scientific coordinator, at, for additional information.

