## Supplementary Material for "Change in body weight of older adults before and during the COVID-19 pandemic: Longitudinal results from the Berlin Aging Study II"

Supplementary Methods

*Study Population*

The interdisciplinary Berlin Aging Study II (BASE-II) aims at the identification of health-promoting factors in old age [1]. The medical part of the baseline examination (T0) included 1,671 older adults (mean age: 68.8 years, sd: 3.7 years, 51.6% women) and was carried out between 2009 and 2014. Between 3.9 and 10.4 years (mean 7.4 years, sd = 1.5 years) later, 1,083 of the originally examined participants took part in the first follow-up examination as part of the GendAge study (T1) [2]. An additional 17 participants were included in T1 which were not medically examined before but were examined at least at one of the other participating BASE-II study sites. The T1 examination period began on June 22nd 2018 and ended on March 10th 2020, shortly before the start of local COVID-19-restrictions. An additional group of 500 younger participants (mean age 28.9 years, sd: 3.1 years) was assessed at baseline, but were not assessed longitudinally to a sufficient extent to be considered in this report. In order to capture the psychological and physiological effects of the COVID-19 pandemic as well as the associated lockdown measures, an online survey was designed and sent to 1365 BASE-II participants who provided their email address in October 2020. Of all participants who were examined at T0 and T1, 540 participants also completed the online survey. For the purpose of this study, one participant was excluded from these longitudinal data because of a probable data entry error and 67 participants were excluded because they did not provide their body weight at one or more of the three time points, at T0, T1, and the online survey, resulting in a final effective study sample of 472 participants providing information (Figure 1B).

As part of the Charité Corona Cross study, a second medical follow-up examination (T2) was conducted between February 2021 and July 2023. Due to our focus on lockdown associated changes, we included only participants who were examined before August 8th, 2022 (T2a) for the purpose of this study. This date was chosen to focus on COVID-19-pandemic associated effects. However, we acknowledge that as a meaningful cut-off date is missing this choice is arbitrary. After applying our exclusion criteria, a sample of 228 participants was available that provided information on all four time-points (T0, T1, online survey, and T2a, see Figure 1B). We want to point out that in this sample four additional participants were excluded because time between online survey and T2a examination was less than 30 days.

The flow chart in Figure 1 provides an overview on how the final datasets were build.

*General characteristics*

*Sex:* Participants were asked to report their sex as part of the self-administered questionnaire as "man" or "women".

*Gender Score:* To separate the effects of gender from effects of sex, the gender score was developed among participants of the BASE-II sample as part of the GendAge study [2]. It incorporates variables reflecting three of the four gender dimensions defined by the WHO: gender roles, gender relations, and institutionalized gender. Lower values indicate more traditionally male characteristics and higher values indicate more traditionally female characteristics [3].

*Cohabitation:* In this study, cohabitation was used to describe whether participants lived alone or not alone. This variable aggregates answers to several questions throughout the BASE-II questionnaire regarding the participants individual living situation. Participants were documented to live "not alone" if they reported to live together with at least one relative, to live in a household with at least two members, lived together with their spouse or lived together with a same-sex partner. Participants who reported to live in a household with 0 household members were reported to live "alone".

*Body Mass Index (BMI)*: BMI at T1 was calculated as measured weight (in kg) divided by squared height (in m). We would like to point out that BMI has been questioned as a measure of obesity in older adults. It is affected by frequently observed age-associated loss of height [4] and therefore does not necessarily accurately reflect changes in body weight. Furthermore, associations between higher BMI and beneficial outcomes in patients with cardiovascular disease (obesity paradox) [5, 6] as well as lower mortality risk [7] were shown before. Therefore, alternative cut-off values for older adults were suggested [8]. However, no consensus on an optimal BMI for older adults exists and we therefore dichotomized BMI based on the cut-off value of 25 m/kg$^2$ which is in line with recommendations of the American Heart Association (AHA)/American College of Cardiology (ACC) [9] and the World Health Organization (WHO) [10].

*Education:* Education was measured in number of years.

*Income:* Participants were asked to report their personal net income within the following ranges: 1) below 300€, 2) 300-599€, 3) 600-999€, 4) 1000-1399, 5) 1400-1799€, 6) 1800-2199€, 7) 2200-1299€, 8) 2500-2999€, 9)3000-3499€, 10) 3500-3999€, 11) 4000-4499€, 12) 4500-4999€, 13) 5000-5499€, 14) 5500-5999€, 15) 6000-6499€, 16) 6500-6999€, 17) 7000-7499€, 18) 7500€ or more.

<u>*Physical Activity*</u>

*Physical activity (PA)*: Participants were asked to rate themselves as “active” or “rarely/never active” (reported as “not active” in this study) as part of the Rapid Assessment of Physical Activity (RAPA) [11] questionnaire.

*Actigraphy (VM counts):* PA was more objectively assessed by the wGT3X-BT activity monitor (ActiGraph LLC, USA). The accelerometer was worn like a wristwatch for on average six days by the BASE-II participants and collected data on acceleration in three axis that was subsequently processed with Actigraphs ActiLife software package. Vector Magnitude (VM) is calculated as square root of the sum of squared acceleration data from each of the three axes. Several other PA estimates like step counts and energy expenditure are available as well, but VM does, in contrast to most other estimates, not rely on cut-offs or algorithms. A detailed description of this variable including its evaluation in the BASE-II cohort can be found in reference[12].

*Change in activity in past 6 months:* To analyze change in PA that could potentially be attributed to the COVID-19 pandemic and associated lockdown measures, participants were asked whether their PA changed within the past six months as part of the online survey (Has your physical activity (e.g., sports activities, walks, etc.) changed due to corona-related limitations in the past 6 months?). Participants were asked to choose between "no change", "PA increased" and "PA decreased". We want to point out that this is the only variable analyzed with respect to differences in body weight change that was assessed during the online survey and not at the first follow-up examination that took place before the Corona outbreak.

*Cognitive performance, Hand Grip Strength and Frailty*

*Digit Symbol Substitution Test (DSST):* Cognitive performance, specifically perceptual speed, of participants was assessed by the standardized DSST. Participants were asked to match symbols to numbers according to a provided key on a single sheet of paper [13].

*Hand Grip Strength:* Muscle strength was assessed as hand grip strength. Participants were asked to perform three maximal isometric contractions with each hand [14]. The highest value of all six contractions was used in the analyses. Impairment was defined by sex- and BMI-stratified cut-off values [15].

*Fried's Frailty Index:* The Frailty index proposed by Linda Fried and colleagues [15] is calculated as a composite marker aggregating unintended weight loss, exhaustion, weakness, slow walking speed, and low physical activity. Each additional impairment in one these variables result in an increase of one point of the final frailty score (possible range: 0 to 5 points). In this study, a score >0 was defined as frail. More information on how the Fried frailty score was determined in BASE-II can be found in reference [14].

*SPRINT-BASEed Frailty Index:* To construct this frailty index we followed the deficit accumulation approach [16] and used 31 of the 37 items from the Systolic Blood Pressure Intervention Trial (SPRINT) described by Pajewsky and colleagues which were available or could be adapted from similar items in BASE-II. The weighting of the individual items, the calculation and cut-offs had also been largely adopted from Pajewski et al., 2016 [17]. As an additional variable we included hand grip strength (with the Fried frailty item "grip strength" cut-offs [15]), so that our SPRINT-BASEed frailty index considered a total of 32 items, of which at least 30 had to be available for an individual to be used in the analyses.

*Cardiovascular Risk and Chronic Disease*

*SCORE2 and SCORE2-OP:* Ten-year risk for cardiovascular disease was calculated as composite score SCORE2 [18]. Older participants (>70 years) risk was calculated with the SCORE2-OP score [19]. As suggested by the authors, SCORE2/SCORE2 OP was not calculated for participants who had a diagnosed diabetes mellitus, myocardial infarction, or stroke.

*Type 2 Diabetes Mellitus (T2D):* T2D was diagnosed based on the criteria defined by the American Diabetes Association (ADA) guidelines [20] as follows: Anamnestic history of diabetes mellitus type 2 (self-report), antidiabetic medication, fasting plasma glucose ≥ 126 mg/dl, 2h plasma glucose during 75g-OGTT ≥ 200mg/dl, and HbA1c ≥ 6.5%. Additional information about T2D in BASE-II can be found in reference [21].

*Metabolic syndrome:*

The metabolic syndrome was diagnosed according to the American Heart Association/ International Diabetes Federation/ National Heart, Lung, and Blood Institute criteria 2009 [22].

*Kidney function*: Kidney function was determined by estimating the glomerular filtration rate (eGFR) using the CAPA equation [23].

*Morbidity Index:* To get a more general estimation of the participants morbidity, an adapted version [24] of Charlson's morbidity index [25] was calculated.

<u>*Biomarkers of Aging*</u>

*Epigenetic Clocks:* To assess the participants biological age, epigenome-based variables were used. Epigenetic clocks allow an estimation of the epigenetic age in years (DNA methylation age, DNAmA). DNAmA was estimated using the 7-CpG clock [26]and GrimAge clock [27] algorithm. In this study we use DNAmA acceleration (DNAmAA) calculated as leukocyte-adjusted residuals of a linear regression of DNAmA on chronological age as measure for each individual's deviation of biological age from chronological age. More information on how DNAmA was measured in BASE-II can be found in reference [28].

*Telomere Length:* Telomeres are the ends of chromosomes and shorten with every replication cycle. Telomere length can be measured directly but a recently available algorithm allows its estimation based on epigenetic data [29].

<u>*Psychological Variables*</u>

*Positive and Negative Affect:* To assess affect, we used the Positive and Negative Affect Schedule (PANAS-X [30]). Internal consistency was acceptable, with Cronbach's α = 0.64 [31].

*Loneliness*: Loneliness was examined using the UCLA Loneliness Scale [32] calculated as a mean score of seven items. The scale contained such items as "There are people I feel close to" or "I feel isolated from others". Participants were asked to rate each statement on a 5-point Likert scale ranging from 1–5 ('1 – strongly disagree' to '5 – strongly agree'). Higher scores indicate stronger feelings of loneliness (Cronbach's α = 0.81) [33].

*Conscientiousness*: To assess the personality disposition of Conscientiousness, we used a 3-item subscale of the short version of the Big Five Inventory (BFI) [34]. Participants were asked to indicate their agreement on a 7-point Likert scale ('1 = does not apply at all' to '7 = applies perfectly'). Internal consistency was modest (Cronbach's $\alpha = 0.63$) [35].

*Cohen's Perceived Stress Scale (PSS):*Items 1, 2, 3, 7, 8, 10, and 11 of the PSS by Cohen and colleagues [36] were used to measure perception of stress. Participants answered the respective questions on a scale from 1 to 5. The results of the individual items were averaged and z-transformed to improve interpretability. Descriptive statistics of the average are shown to increase comparability to other studies in Supplementary Table 1. Normalized values were used in the imputation process and subsequent analyses. Additional information on the PSS in BASE-II can be found in reference [37].

*Missing Values*

Multiple imputations were carried out under the missing at random assumption. Number of observed/missing values in each variable is shown in Supplementary Table 3. 5.2% of the data was missing and subsequently imputed. Of all analyzed participants, 139 had a complete dataset and 293 participants had missing values in 1 to 3 variables. Only 40 participants had missing values in more than 3 variables. No individual combination of two or more missing values appeared more than 14 times. Multiple imputations were done with R's mice function (mice package [38]) generating 20 imputed datasets over 5 iterations (m=20, maxit=5). Mice's predictive mean matching method ("pmm") was used which allows the imputation of discrete variables. Because only values that were already observed in the dataset are used for imputation, no meaningless values or values more extreme than those already present in the originally observed data are present in the imputed datasets.

All variables of interest in this study (which are part of Supplementary Table 1) as well as the outcome variable (weight CAGR between T1 and online survey and others) and additional variables (Horvath DNAmAA, Hannum DNAmAA, PhenoAge DNAmAA, CKD-EPI GFR and others) were included in the imputation procedure. As no statistical interaction terms were analyzed in this study, we refrained from including them in the imputation process. Convergence of the imputation algorithm was inspected visually by plotting summaries of the imputed values across the iterations. Distribution and proportions of imputed variables were compared with the observed data visually as density plots and bar plots and were consistent across datasets.

To ensure internal consistency of the data, dichotomization (if necessary) was performed after the imputation in each dataset individually. In case of cut-off values defined by the median, distributions of each of the imputed dataset were used individually to define cut-off values. Therefore, number of participants per groups as well as cut-off values can potentially vary between the 20 imputed datasets. After the dichotomization, datasets were combined by the "imputationList" function (mitools package [39]).

In follow-up analyses, we re-ran our models using only originally observed and not-imputed data. Results are reported in Supplementary Figure 5 and 6 and revealed a highly comparable pattern of results to those reported in the main text.

*Statistical Analyses*

All statistical analyses were conducted in R, version 4.2.1 [40]. Figures were drawn with R's "ggplot2" package [41]. Forest plots were constructed with the "forestplot" package [42]. The package tableone [43] was used to build the descriptive statistics tables. This package employs the definition of standardized mean difference as described by Flury and Riedwyl [44] in 1986. Mean difference between groups as well as 95% confidence intervals and p-values for two-sided two-sample t-tests were calculated and pooled across imputed datasets with the mi.t.test function (MKmisc package [45]). In the not-imputed dataset, mean and 95% confidence intervals were calculated with the *groupwiseMean* function (rcompanion package [46]). The regression line equation, Pearson's correlation coefficient and $R^2$ were calculated with the *stat_regline_equation* and *stat_cor* function of the ggpubr package [47]. The alluvial plot was drawn with help of the ggalluvial package [48]. Statistical significance was set at a level of 5%.

We chose to dichotomize continuous variables whenever possible and used dummy variables if binary categorization was not an option. This was done to present easy to interpret results for all variables in the same way. We acknowledge that this methodological approach is associated with loss of information and possibly statistical power [49] but is necessary to allow an easy interpretation and comparison of results between variables. Additionally, a correlation plot of continuously measured variables at T1 and weight change between T1 and the online survey did not reveal any differing results (Supplementary Figure 7).

Supplementary Material

Tables

Supplementary Table 1: Descriptive statistics of analyzed variables at T1 allowing a comparison between BASE-II participants that provided data in the Online Survey and are included in the current study and participants that did not participate in the Online Survey.

| | | | Followed-up in Online Survey | | | | | Not Followed-up in Online Survey | | | | | | |
|---|---|---|---|---|---|---|---|---|---|---|---|---|---|---|
| | Variable (at T1) | Category | n | mean, % | sd | min | max | n | mean | sd | min | max | SMD | p-value |
| General Characteristics | Age | | 472 | 74.82 | 3.65 | 64.91 | 85.94 | 628 | 76.20 | 3.75 | 66.99 | 94.07 | 0.374 | 0.000 |
| | Sex | Men | 245 | 51.91 | | | | 282 | 44.90 | | | | | 0.025 |
| | | Women | 227 | 48.09 | | | | 346 | 55.10 | | | | | |
| | Genderscore | | 387 | 0.50 | 0.34 | 0.00 | 1.00 | 512 | 0.54 | 0.35 | 0.00 | 1.00 | 0.123 | 0.068 |
| | Cohabitation | live not alone | 297 | 69.23 | | | | 350 | 64.10 | | | | | 0.107 |
| | | live alone | 132 | 30.77 | | | | 196 | 35.90 | | | | | |
| | BMI | | 472 | 26.78 | 3.98 | 17.17 | 49.26 | 626 | 27.12 | 4.44 | 17.85 | 49.68 | 0.081 | 0.187 |
| | Education (years) | | 451 | 14.77 | 2.85 | 7.00 | 18.00 | 543 | 14.14 | 2.94 | 7.00 | 18.00 | 0.218 | 0.001 |
| | Income | | 455 | 6.87 | 4.02 | 1.00 | 20.00 | 610 | 6.43 | 3.99 | 1.00 | 20.00 | 0.110 | 0.077 |
| Physical Activity | Actigraphy (VM counts x100,000) | | 345 | 195.90 | 53.13 | 46.04 | 355.30 | 415 | 190.80 | 56.12 | 46.41 | 438.30 | 0.094 | 0.199 |
| | Physical Activity (self-assessed) | active | 408 | 86.62 | | | | 556 | 88.96 | | | | | 0.279 |
| | | not active | 63 | 13.38 | | | | 69 | 11.04 | | | | | |
| Frailty, HGS, cognitive | SPRINT BASEed Frailty Index | 1 | 107 | 24.04 | | | | 108 | 18.46 | | | | | 0.000 |
| | | 2 | 276 | 62.02 | | | | 313 | 53.50 | | | | | |
| | | 3 | 62 | 13.93 | | | | 164 | 28.03 | | | | | |
| | Fried Frailty score | | 467 | 0.62 | 0.80 | 0.00 | 4.00 | 620 | 0.87 | 0.91 | 0.00 | 4.00 | 0.286 | 0.000 |

| | Variable (at T1) | Category | Followed-up in Online Survey n | mean, % | sd | min | max | Not Followed-up in Online Survey n | mean | sd | min | max | SMD | p-value |
|---|---|---|---|---|---|---|---|---|---|---|---|---|---|---|
| | Hand grip strength | | 472 | 28.73 | 9.31 | 7.50 | 57.00 | 626 | 26.50 | 9.06 | 6.00 | 56.00 | 0.243 | 0.000 |
| | DSST | | 471 | 42.32 | 8.07 | 2.00 | 72.00 | 624 | 39.61 | 8.93 | 4.00 | 73.00 | 0.318 | 0.000 |
| Chronic Disease | Morbidity Index | | 413 | 1.42 | 1.52 | 0.00 | 8.00 | 541 | 1.38 | 1.56 | 0.00 | 9.00 | 0.028 | 0.665 |
| | eGFR Cystatin C | | 470 | 74.89 | 15.17 | 7.50 | 90.00 | 623 | 71.28 | 16.16 | 19.60 | 90.00 | 0.231 | 0.000 |
| | SCORE2/SCORE2 OP | | 362 | 15.00 | 5.38 | 5.00 | 36.00 | 481 | 16.35 | 6.15 | 5.00 | 41.00 | 0.235 | 0.001 |
| | Metabolic Syndrom | no | 242 | 52.38 | | | | 342 | 55.43 | | | | | 0.351 |
| | | yes | 220 | 47.62 | | | | 275 | 44.57 | | | | | |
| | Diabetes Mellitus Type 2 | no DM diagnosed | 387 | 81.99 | | | | 509 | 83.58 | | | | | 0.544 |
| | | DM diagnosed | 85 | 18.01 | | | | 100 | 16.42 | | | | | |
| Biomar kerks of | 7-CpG IEAA | | 460 | -0.33 | 4.94 | -15.00 | 25.06 | 603 | 0.27 | 5.21 | -16.97 | 19.42 | 0.119 | 0.056 |
| | GrimAge IEAA | | 460 | 0.11 | 3.45 | -10.91 | 11.05 | 603 | -0.03 | 3.33 | -10.05 | 12.84 | 0.040 | 0.513 |
| | DNAmTL | | 467 | 6.82 | 0.19 | 6.11 | 7.37 | 612 | 6.79 | 0.20 | 5.98 | 7.28 | 0.188 | 0.002 |
| Psychosocial | Positive Affect | | 439 | 3.91 | 0.57 | 2.00 | 5.00 | 557 | 3.80 | 0.62 | 1.40 | 5.00 | 0.186 | 0.004 |
| | Negative Affect | | 442 | 2.24 | 0.70 | 1.00 | 4.50 | 556 | 2.36 | 0.73 | 1.00 | 4.75 | 0.171 | 0.008 |
| | Loneliness | | 445 | 1.56 | 0.59 | 1.00 | 4.43 | 553 | 1.65 | 0.67 | 1.00 | 4.57 | 0.142 | 0.026 |
| | Conscientiousness | | 443 | 5.62 | 0.98 | 2.33 | 7.00 | 559 | 5.57 | 1.01 | 1.67 | 7.00 | 0.048 | 0.453 |
| | Cohen's PSS | | 445 | 2.00 | 0,62 | 1 | 4.38 | 561 | 2.14 | 0.65 | 1 | 4.5 | 0.214 | 0.001 |

Note: sd = Standard Deviation, Min = Minimum, Max = Maximum, SMD = Standardized Mean Difference, IEAA = intrinsic epigenetic age acceleration.

Supplementary Table 2: Overview of analyzed variables and how they were dichotomized if needed. Additional information on methods and how they were assessed are provided in the last column.

| **Category** | **Variable** | **Grouping methods**<br>**[group1; group 2]**<br>**Further information** | **References for further information:** |
|---|---|---|---|
| General Characteristics | Sex | Dichotomous Variable<br>[“men”; “women”] | |
| | Gender Score | Median Split<br>[“below/equal median”; “above median”]<br>The gender score was developed to separate the effects of gender from sex in older adults. It was developed in the sample of 1100 participants of the GendAge study. | [3] |
| | Cohabitation | Dichotomous Variable<br>[“live alone”; “live not alone”]<br>Cohabitation was derived by aggregating answers to several questions distributed over the questionnaires employed. This included answers to the question “What is your relationship to the other persons in your household?” and whether they live together or in separation with their partner. | n/a |
| | BMI | Cut-off-based<br>[<25; >=25]<br>Cut-off based on recommendations of American Heart Association (AHA)/American College of Cardiology (ACC) [9] and the World Health Organization (WHO) [10] | [1, 2] |
| | Education | Median Split<br>[”below median”, “above/equal median”]<br>Total education years were assessed by participants self-report. | |
| | Income | Median Split<br>[”below median”, “above/equal median”]<br>Participants reported their individual net income in one the following categories: 1) below 300€, 2) 300-599€, 3) 600-999€, 4) 1000-1399, 5) 1400-1799€, 6) 1800-2199€, 7) 2200-1299€, 8) 2500-2999€, 9)3000-3499€, 10) 3500-3999€, 11) 4000-4499€, 12) 4500-4999€, 13) 5000-5499€, 14) 5500-5999€, 15) 6000-6499€, 16) 6500-6999€, 17) 7000-7499€, 18) 7500€ or more. | |
| Physical Activity | Physically active (self-reported) | Dichotomous Variable<br>[“active”; “not active”]<br>Answer to Question 1 of Rapid Assessment of Physical Activity (RAPA) [11] | [2] |
| | Actigraphy (VM counts) | Median Split<br>[“below/equal median”; “above median”]<br>VM calculated as square root of the sum of squared data from each of the three axis measured by an Actigraph wGT3X-BT accelerometer. | [50] |
| | Change in activity in past 6 months | Categorical Variable<br>[“activity reduced”; “no change”; “activity increased”] | n/a |

| | | | |
|---|---|---|---|
| | | Participants were asked how their PA changed over the past 6 months during the online survey. Possible answers were "no change", "PA increased" and "PA decreased". Dichotomization was not possible, therefore "no change" was used as reference category. | |
| Cognitive assessments, Hand Grip Strength and Frailty | DSST [13] | Median Split<br>["below/equal median"; "above median"]<br>This paper-and-pencil test asks participants to match symbols to numbers according to a given key and measures cognitive performance. | [51] |
| | Hand Grip Strength | Cut-off-based<br>[not impaired; impaired]<br>Cut-offs stratified by sex and BMI and defined by Fried and colleagues were used [15]:<br><br>Men:<br>BMI<=24 & HGS<=29 or 24>BMI>=26 & HGS<=30 or 26<BMI<=28 & HGS<=30 oder BMI>28 & HGS<=23<br><br>Women:<br>BMI<=23 & HGS <=17 or 23>BMI<=26 & HGS<=17.3 or 26<BMI<29 & HGS<=18 or BMI>29 & HGS <=21 | [14] |
| | Fried's Frailty phenotype [15] | Cut-off-based<br>[not frail; frail]<br>Participants with a frailty score higher than 0 were considered frail [14]. | [14] |
| | SPRINT-BASEed Frailty Index | Cut-of-based<br>["fit"; "less fit"; "frail"]<br>This frailty index was calculated based on the algorithm published by Pajewski et al., 2016. [52] | n/a |
| Chronic Disease | SCORE2 and SCORE2-OP[18, 19] | Median split<br>[below/ equal median; above median]<br>Participants were categorized by median split. | |
| | eGFR (Cystatin C) | Cut-off based<br>[normal kidney function; reduced kidney function]<br>Reduced kidney function was defined as a GFR <60 mL/min/1.73 m$^2$ [53], | [53] |
| | Type 2 Diabetes Mellitus | Dichotomous Variable<br>[no T2D diagnosed; T2D diagnosed]<br>T2D was diagnosed following the American Diabetes Association (ADA) guidelines [20]. | [21] |
| | Metabolic Syndrome [22] | Dichotomous Variable<br>[no; yes]<br>The Metabolic Syndrome was diagnosed according to the American Heart Association/ International Diabetes Federation/ National Heart, Lung,and Blood Institute criteria 2009 . [2] | |
| | Morbidity Index [25] | Cut-off-based<br>[0; 1-2; >2] | [24] |

| | | | |
|---|---|---|---|
| | | An adapted version of the Morbidity Index by Charlson and colleagues [25] was used in is described in detail elsewhere [24]. To allow a better evaluation of the general morbidity burden participants were grouped. Cut-off were chosen arbitrarily to allow comparison between groups. | |
| Biomarkers of Aging | DNAmAA [26, 27, 54-56] | Cut-off-based<br>[biologically younger; biologically older]<br>Participants with DNAmAA <0 were categorized as biologically younger. Participants with DNAmAA >0 were categorized as biologically older. | [26, 28] |
| | DNAmTL [29] | Median Split<br>[“below/equal median”; “above median”]<br>Participants were categorized based on a median split. | |
| Psychosocial | Positive Affect | Median Split<br>[“below/equal median”; “above median”]<br>Affect was assessed by the Positive and Negative Affect Schedule (PANAS-X [30]). | [31] |
| | Negative Affect | Median Split<br>[“below/equal median”; “above median”]<br>Affect was assessed by the Positive and Negative Affect Schedule (PANAS-X [30]). | [31] |
| | Loneliness | Median Split<br>[“below/equal median”; “above median”]<br>Loneliness was assessed with the UCLA Loneliness Scale [32]. | [33] |
| | Conscientiousness | Median Split<br>[“below/equal median”; “above median”]<br>Assessed with 3-item subscale of the short version of the Big Five Inventory (BFI) [34]. | [35] |
| | Cohen’s Perceived Psychological Stress Scale | Meidan Split<br>[“below median”; “above/equal median”]<br>Perception of stress was measured by eight items of the Perceived Stress Scale (PSS) [36]. | [37] |

Supplementary Table 2: Descriptive statistics of age and body weight at the four available timepoints, all available cases are included.

| | | Women | | | | | Men | | | | | |
|---|---|---|---|---|---|---|---|---|---|---|---|---|
| Timepoint | Variable | n | mean | sd | min | max | n | mean | sd | min | max | p-value |
| T0 | Age (years) | 227 | 67.30 | 2.93 | 61.30 | 75.60 | 245 | 67.69 | 3.52 | 60.20 | 77.30 | 0.186 |
| | Body weight (kg) | 227 | 69.22 | 12.16 | 46.50 | 127.00 | 245 | 82.85 | 10.58 | 60.50 | 130.00 | 0.000 |
| T1 | Age (years) | 227 | 74.94 | 3.18 | 66.40 | 85.40 | 245 | 74.70 | 4.04 | 64.90 | 85.90 | 0.468 |
| | Body weight (kg) | 227 | 69.07 | 12.33 | 43.00 | 130.00 | 245 | 81.80 | 10.83 | 60.00 | 125.00 | 0.000 |
| Online Survey | Age (years) | 227 | 76.46 | 3.13 | 68.60 | 86.30 | 245 | 76.22 | 4.01 | 67.20 | 87.70 | 0.459 |
| | Body weight (kg) | 227 | 68.25 | 12.75 | 45.00 | 130.50 | 245 | 81.19 | 10.40 | 60.00 | 125.00 | 0.000 |
| T2a | Age (years) | 111 | 77.40 | 2.98 | 70.20 | 85.20 | 117 | 77.37 | 3.59 | 69.80 | 88.80 | 0.952 |
| | Body weight (kg) | 111 | 66.24 | 11.39 | 43.00 | 103.00 | 117 | 80.40 | 11.01 | 61.00 | 130.00 | 0.000 |
| T0 to T1 | Body weight difference (kg) | 227 | -0.15 | 3.99 | -18.00 | 14.00 | 245 | -1.05 | 5.12 | -21.50 | 15.50 | 0.034 |
| | Body weight difference per year (kg/year) | 227 | -0.03 | 0.57 | -3.10 | 2.60 | 245 | -0.13 | 0.73 | -3.20 | 2.50 | 0.082 |
| | Body weight change CAGR (%) | 227 | -0.04 | 0.82 | -3.30 | 4.50 | 245 | -0.17 | 0.86 | -3.30 | 3.20 | 0.100 |
| T1 to Online | Body weight difference (kg) | 227 | -0.82 | 3.21 | -14.00 | 11.10 | 245 | -0.61 | 3.57 | -21.50 | 13.00 | 0.512 |
| Survey | Body weight difference per year (kg/year) | 227 | -0.57 | 2.20 | -11.50 | 6.60 | 245 | -0.48 | 2.46 | -16.10 | 12.90 | 0.678 |
| | Body weight change CAGR (%) | 227 | -0.87 | 3.19 | -17.00 | 9.40 | 245 | -0.50 | 2.99 | -17.70 | 21.80 | 0.203 |
| Online Survey | Body weight difference (kg) | 111 | -0.06 | 2.74 | -14.00 | 10.00 | 117 | 0.28 | 3.76 | -13.00 | 20.00 | 0.446 |
| to T2a | Body weight difference per year (kg/year) | 111 | -0.05 | 2.15 | -10.30 | 7.80 | 117 | 0.66 | 4.88 | -8.20 | 43.20 | 0.159 |
| | Weight change CAGR (%) | 111 | -0.02 | 3.10 | -13.80 | 12.10 | 117 | 0.93 | 7.37 | -8.60 | 72.10 | 0.212 |

Figures

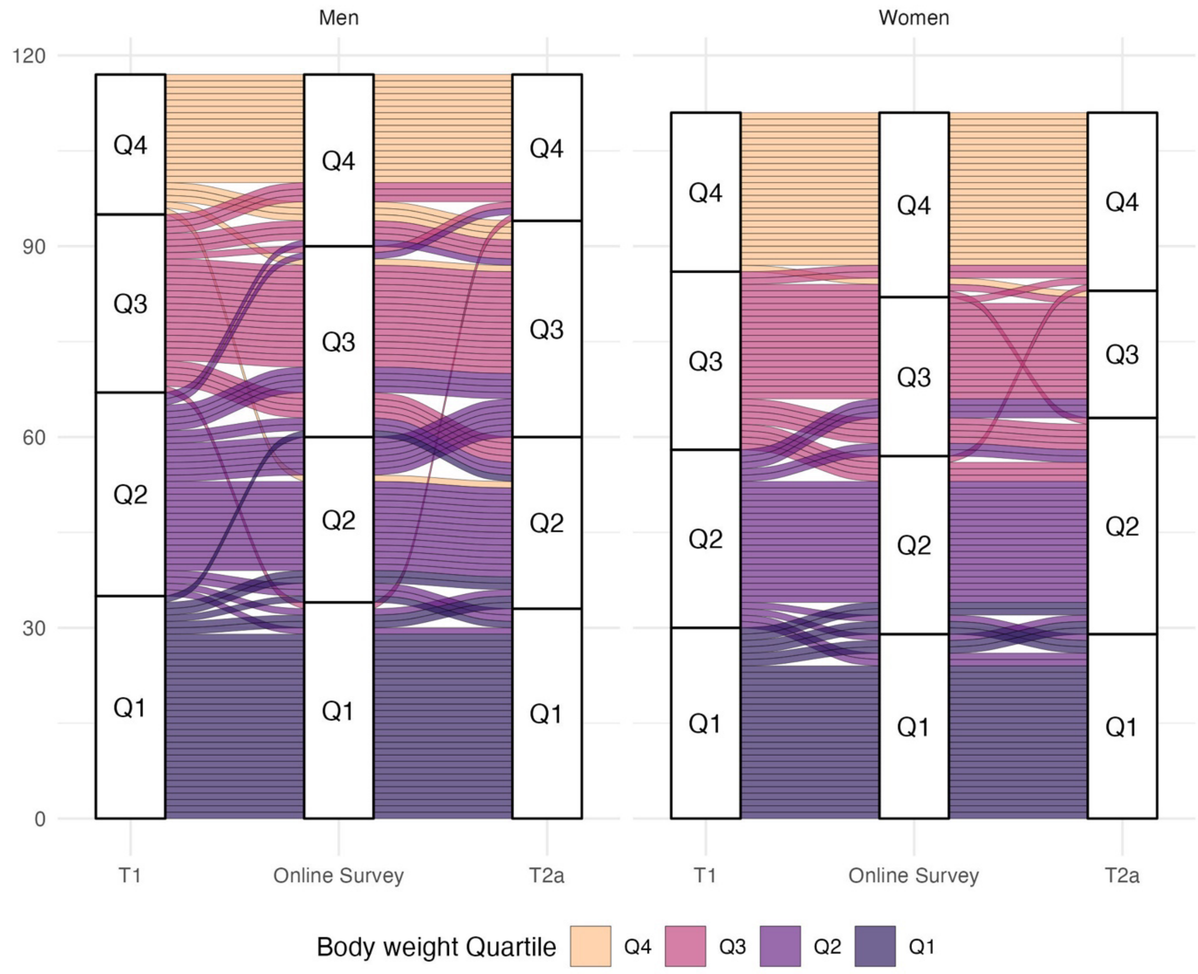


Supplementary Figure 1: Alluvial plot of weight quartiles of the core sample (n=227) at T1, Online Survey and T2a.

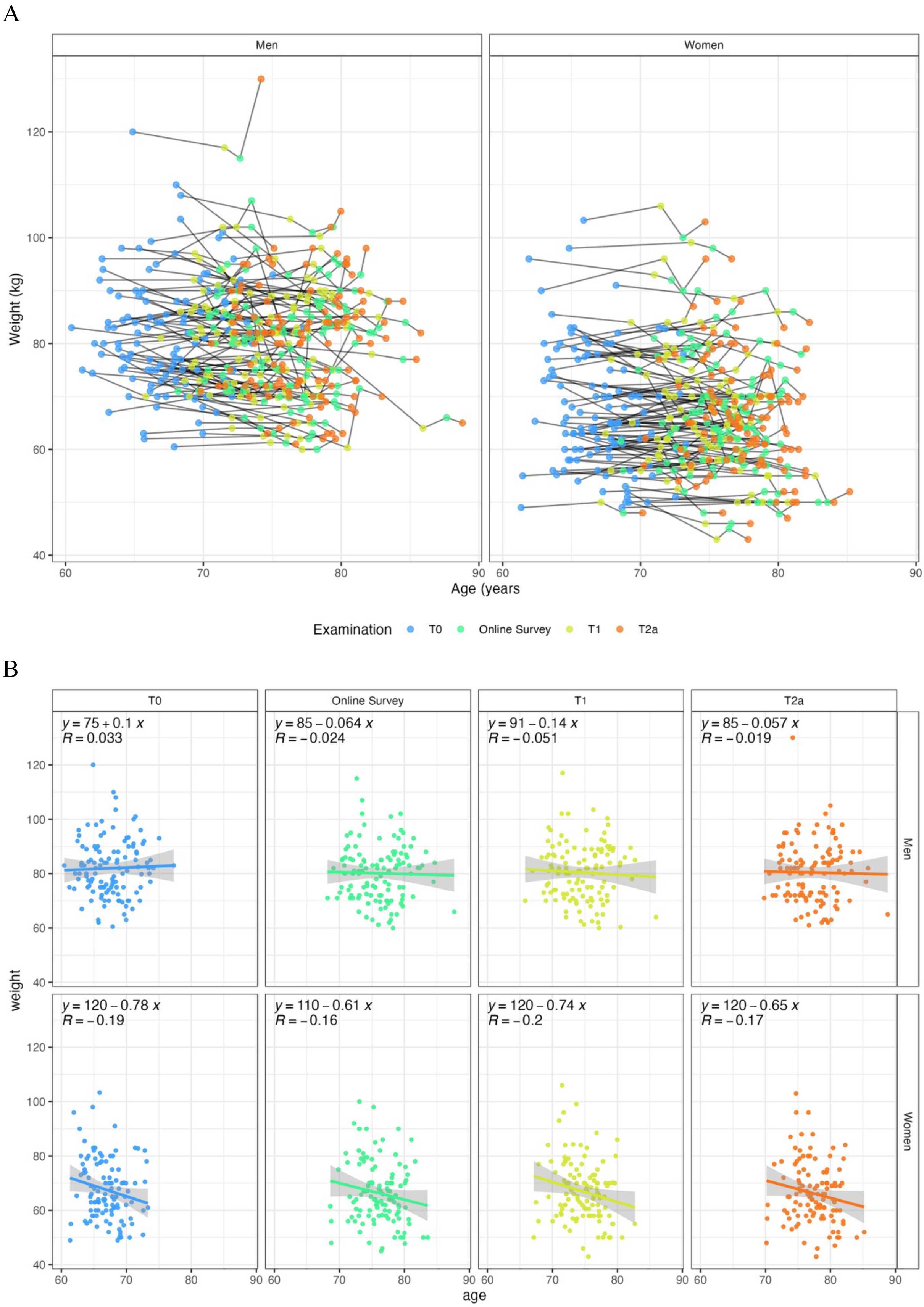


Supplementary Figure 2: A: individual trajectories of participants over the course of four examinations (n=227) stratified by sex. B: Scatterplots of weight and age at four time points stratified by sex (n=227).

### Women And Men

| Variable | Group | Mean | SD | Group | Mean | SD | p-value |
|---|---|---|---|---|---|---|---|
| Cohen's Perceived Stress Scale | below median | -0.91 | 2.86 | above/equal median | -0.49 | 3.37 | 0.157 |
| Gender Score | below/equal median | -0.85 | 3.15 | above median | -0.5 | 3.17 | 0.239 |
| Negative Affect | below/equal median | -0.81 | 3.19 | above median | -0.48 | 3.04 | 0.263 |
| DSST | below/equal median | -0.83 | 3.47 | above median | -0.53 | 2.66 | 0.298 |
| Positive Affect | below median | -0.81 | 3.11 | above/equal median | -0.55 | 3.25 | 0.385 |
| Income | below median | -0.8 | 3.05 | above/equal median | -0.57 | 3.24 | 0.432 |
| GrimAge DNAmAA | biologically younger | -0.72 | 2.82 | biologically older | -0.63 | 3.37 | 0.75 |
| Cohabitation | live not alone | -0.7 | 3.19 | live alone | -0.64 | 3.21 | 0.855 |
| FI SPRINT BASEed | fit | -0.62 | 2.45 | frail | -0.6 | 3.29 | 0.957 |
| FI Fried | not frail | -0.68 | 3.24 | frail | -0.67 | 2.91 | 0.972 |
| Physically active | active | -0.68 | 3.07 | not active | -0.67 | 3.26 | 0.984 |
| Handgrip Strength | impaired | -0.69 | 2.79 | not impaired | -0.68 | 3.18 | 0.973 |
| Loneliness | below median | -0.67 | 2.72 | above/equal median | -0.68 | 3.55 | 0.971 |
| BMI | < 25 | -0.67 | 3.48 | >= 25 | -0.68 | 2.86 | 0.968 |
| FI SPRINT BASEed | fit | -0.62 | 2.45 | less fit | -0.72 | 3.31 | 0.765 |
| DNAmTL | below/equal median | -0.61 | 2.95 | above median | -0.75 | 3.23 | 0.626 |
| SCORE2 | below/equal median | -0.62 | 3.17 | above median | -0.76 | 3.41 | 0.665 |
| Conscientiousness | below/equal median | -0.59 | 2.93 | above median | -0.78 | 3.37 | 0.521 |
| Education | below median | -0.58 | 2.62 | above/equal median | -0.77 | 3.52 | 0.516 |
| Actigraphy (VM counts) | below/equal median | -0.56 | 3.57 | above median | -0.8 | 3.31 | 0.483 |
| Sex | 1 | -0.5 | 2.99 | 2 | -0.87 | 3.19 | 0.203 |
| 7-CpG DNAmAA | biologically younger | -0.5 | 3.34 | biologically older | -0.88 | 2.82 | 0.189 |
| MetS | no | -0.47 | 3.01 | yes | -0.91 | 3.24 | 0.137 |
| Morbidity Index | 0 | -0.3 | 3.34 | 1-2 | -0.74 | 2.83 | 0.177 |
| Diabetes Mellitus | no DM diagnosed | -0.58 | 2.95 | DM diagnosed | -1.12 | 3.63 | 0.147 |
| GFR | normal kidney function | -0.58 | 2.78 | reduced kidney function | -1.12 | 4.19 | 0.139 |
| Morbidity Index | 0 | -0.3 | 3.34 | >2 | -1.19 | 3.44 | 0.042 |

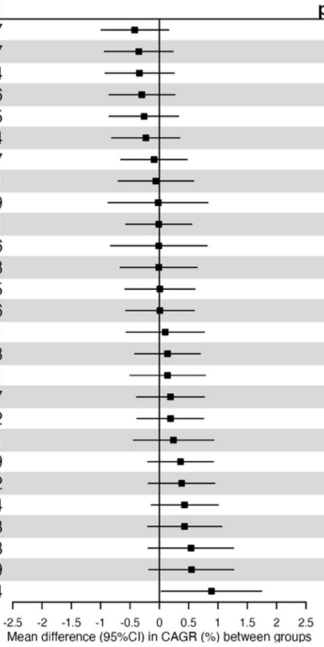


Supplementary Figure 3: Forest plot of mean difference and 95% CI in CAGR (%) between groups in the imputed dataset of women and men.

### Women and Men (not-imputed)

| Variable | Group | Mean | n | Group | Mean | n |
|---|---|---|---|---|---|---|
| Cohen's Perceived Stress Scale | below median | -0.834 | 191 | above/equal median | -0.435 | 254 |
| Negative Affect | below/equal median | -0.754 | 267 | above median | -0.366 | 175 |
| DSST | below/equal median | -0.836 | 236 | above median | -0.541 | 235 |
| Gender Score | below/equal median | -0.745 | 194 | above median | -0.473 | 193 |
| Income | below median | -0.794 | 216 | above/equal median | -0.546 | 239 |
| Positive Affect | below median | -0.703 | 213 | above/equal median | -0.489 | 226 |
| GrimAge DNAmAA | biologically younger | -0.763 | 230 | biologically older | -0.655 | 230 |
| FI SPRINT BASEed | fit | -0.699 | 107 | frail | -0.599 | 62 |
| Cohabitation | live not alone | -0.641 | 297 | live alone | -0.573 | 132 |
| FI Fried | not frail | -0.665 | 252 | frail | -0.646 | 215 |
| Loneliness | below median | -0.617 | 207 | above/equal median | -0.598 | 238 |
| Handgrip Strength | impaired | -0.687 | 113 | not impaired | -0.676 | 359 |
| Physically active | active | -0.677 | 408 | not active | -0.666 | 63 |
| BMI | < 25 | -0.671 | 167 | >= 25 | -0.683 | 305 |
| SCORE2 | below/equal median | -0.623 | 183 | above median | -0.669 | 179 |
| FI SPRINT BASEed | fit | -0.699 | 107 | less fit | -0.755 | 276 |
| Conscientiousness | below/equal median | -0.521 | 229 | above median | -0.655 | 214 |
| DNAmTL | below/equal median | -0.607 | 234 | above median | -0.745 | 233 |
| Education | below median | -0.528 | 217 | above/equal median | -0.731 | 234 |
| Actigraphy (VM counts) | below/equal median | -0.557 | 173 | above median | -0.849 | 172 |
| Sex | 1 | -0.504 | 245 | 2 | -0.867 | 227 |
| 7-CpG DNAmAA | biologically younger | -0.536 | 244 | biologically older | -0.905 | 216 |
| MetS | no | -0.528 | 242 | yes | -0.934 | 220 |
| Morbidity Index | 0 | -0.237 | 146 | 1-2 | -0.758 | 184 |
| Diabetes Mellitus | no DM diagnosed | -0.582 | 387 | DM diagnosed | -1.12 | 85 |
| GFR | normal kidney function | -0.585 | 384 | reduced kidney function | -1.12 | 86 |
| Morbidity Index | 0 | -0.237 | 146 | >2 | -1.38 | 83 |

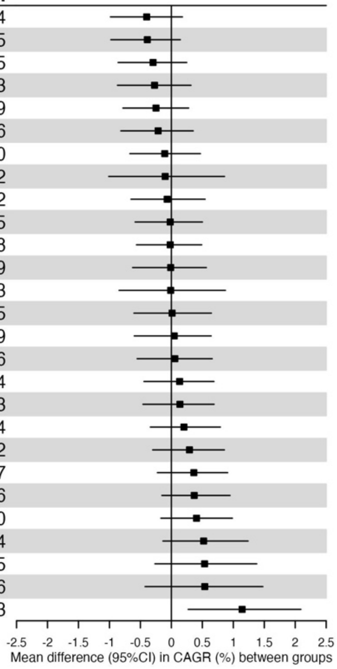


Supplementary Figure 4: Forest plot of mean difference and 95% CI between groups in original (not-imputed) dataset of women and men.

**Women (not-imputed)**

| Variable | Group | Mean | n | Group | Mean | n |
|---|---|---|---|---|---|---|
| Physically active | active | -0.982 | 197 | not active | -0.0481 | 29 |
| Cohen's Perceived Stress Scale | below median | -1.25 | 95 | above/equal median | -0.406 | 118 |
| Gender Score | below/equal median | -1.54 | 40 | above median | -0.733 | 148 |
| FI SPRINT BASEed | fit | -0.844 | 47 | frail | -0.0518 | 39 |
| BMI | < 25 | -1.13 | 98 | >= 25 | -0.666 | 129 |
| GrimAge DNAmAA | biologically younger | -1.02 | 150 | biologically older | -0.74 | 74 |
| DSST | below/equal median | -1.02 | 103 | above median | -0.776 | 123 |
| Negative Affect | below/equal median | -0.897 | 121 | above median | -0.651 | 90 |
| FI Fried | not frail | -0.948 | 117 | frail | -0.739 | 107 |
| Income | below median | -0.95 | 120 | above/equal median | -0.766 | 103 |
| Handgrip Strength | impaired | -0.933 | 59 | not impaired | -0.844 | 168 |
| 7-CpG DNAmAA | biologically younger | -0.946 | 121 | biologically older | -0.909 | 103 |
| DNAmTL | below/equal median | -0.871 | 96 | above median | -0.864 | 131 |
| Actigraphy (VM counts) | below/equal median | -0.965 | 59 | above median | -0.993 | 109 |
| Cohabitation | live not alone | -0.692 | 101 | live alone | -0.839 | 96 |
| MetS | no | -0.893 | 137 | yes | -1.04 | 83 |
| Education | below median | -0.734 | 114 | above/equal median | -0.977 | 106 |
| SCORE2 | below/equal median | -0.76 | 125 | above median | -1.04 | 63 |
| FI SPRINT BASEed | fit | -0.844 | 47 | less fit | -1.15 | 133 |
| Loneliness | below median | -0.594 | 104 | above/equal median | -0.959 | 109 |
| Conscientiousness | below/equal median | -0.541 | 95 | above median | -0.92 | 117 |
| Positive Affect | below median | -0.557 | 97 | above/equal median | -0.994 | 113 |
| Morbidity Index | 0 | -0.335 | 71 | 1-2 | -0.825 | 84 |
| GFR | normal kidney function | -0.747 | 198 | reduced kidney function | -1.68 | 29 |
| Diabetes Mellitus | no DM diagnosed | -0.749 | 201 | DM diagnosed | -1.78 | 26 |
| Morbidity Index | 0 | -0.335 | 71 | >2 | -1.79 | 40 |

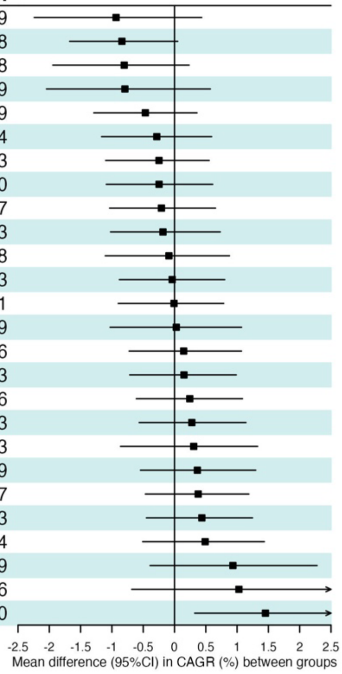


Supplementary Figure 5: Forest plot of mean difference and 95% CI between groups in original (not-imputed) dataset of women only.

**Men (not-imputed)**

| Variable | Group | Mean | n | Group | Mean | n |
|---|---|---|---|---|---|---|
| Gender Score | below/equal median | -0.539 | 154 | above median | 0.383 | 45 |
| Positive Affect | below median | -0.826 | 116 | above/equal median | 0.015 | 113 |
| Cohabitation | live not alone | -0.615 | 196 | live alone | 0.137 | 36 |
| Negative Affect | below/equal median | -0.635 | 146 | above median | -0.064 | 85 |
| DSST | below/equal median | -0.691 | 133 | above median | -0.282 | 112 |
| Loneliness | below median | -0.641 | 103 | above/equal median | -0.293 | 129 |
| Income | below median | -0.599 | 96 | above/equal median | -0.38 | 136 |
| FI SPRINT BASEed | fit | -0.585 | 60 | less fit | -0.387 | 143 |
| Conscientiousness | below/equal median | -0.507 | 134 | above median | -0.335 | 97 |
| Cohen's Perceived Stress Scale | below median | -0.426 | 96 | above/equal median | -0.461 | 136 |
| Handgrip Strength | impaired | -0.419 | 54 | not impaired | -0.528 | 191 |
| FI Fried | not frail | -0.42 | 135 | frail | -0.554 | 108 |
| SCORE2 | below/equal median | -0.328 | 58 | above median | -0.469 | 116 |
| DNAmTL | below/equal median | -0.423 | 138 | above median | -0.593 | 102 |
| Education | below median | -0.3 | 103 | above/equal median | -0.527 | 128 |
| Actigraphy (VM counts) | below/equal median | -0.346 | 114 | above median | -0.6 | 63 |
| GrimAge DNAmAA | biologically younger | -0.275 | 80 | biologically older | -0.615 | 156 |
| Diabetes Mellitus | no DM diagnosed | -0.401 | 186 | DM diagnosed | -0.828 | 59 |
| GFR | normal kidney function | -0.412 | 186 | reduced kidney function | -0.842 | 57 |
| Morbidity Index | 0 | -0.145 | 75 | 1-2 | -0.703 | 100 |
| BMI | < 25 | -0.0157 | 69 | >= 25 | -0.695 | 176 |
| 7-CpG DNAmAA | biologically younger | -0.132 | 123 | biologically older | -0.9 | 113 |
| Physically active | active | -0.393 | 211 | not active | -1.19 | 34 |
| MetS | no | -0.0515 | 105 | yes | -0.867 | 137 |
| Morbidity Index | 0 | -0.145 | 75 | >2 | -1 | 43 |
| FI SPRINT BASEed | fit | -0.585 | 60 | frail | -1.53 | 23 |

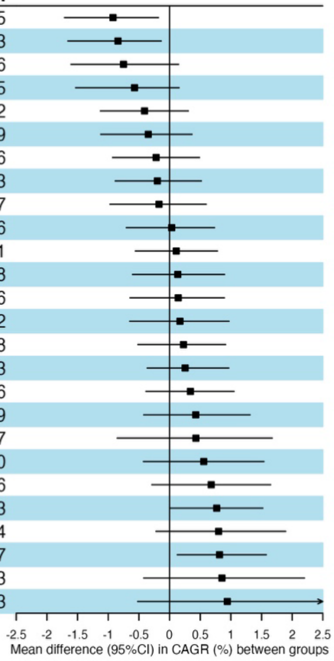


Supplementary Figure 6: Forest plot of mean difference and 95% CI between groups in original (not-imputed) dataset of men only.

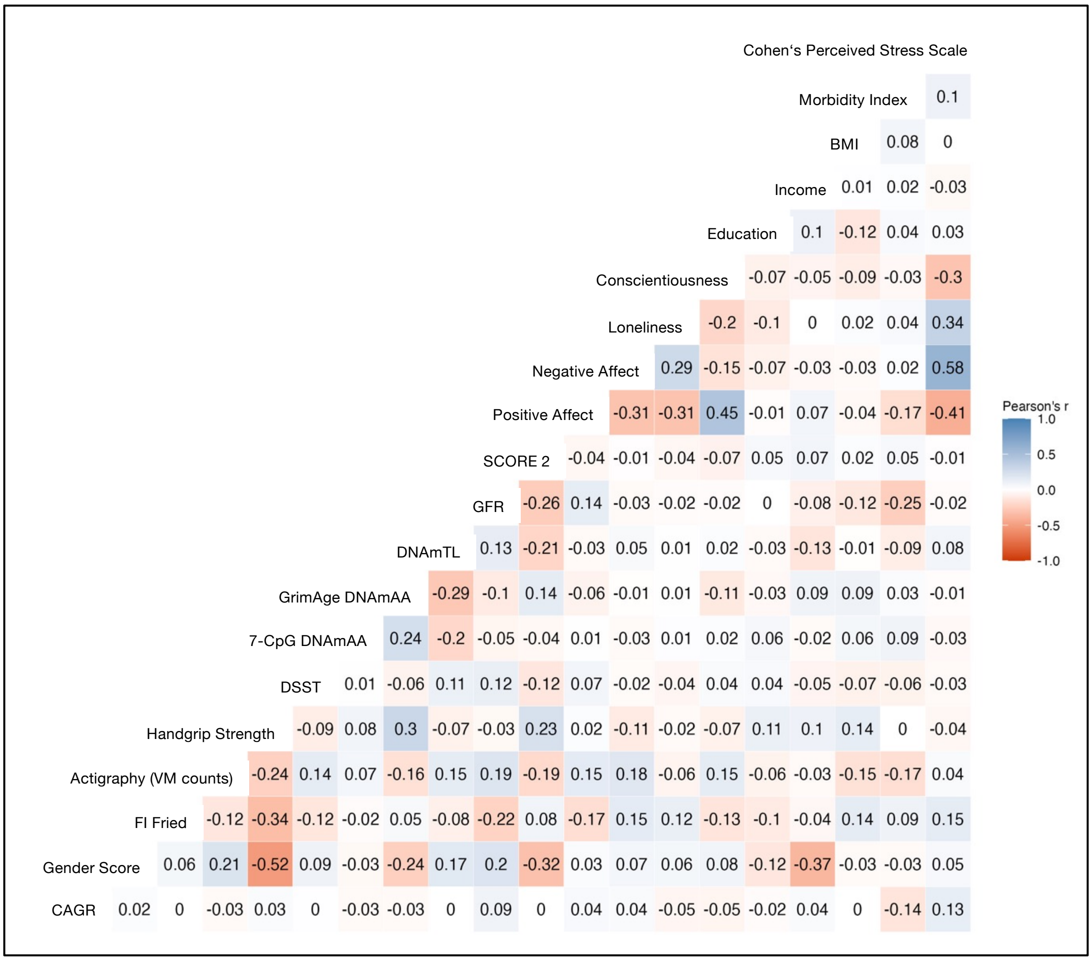


Supplementary Figure 7: Correlation Plot of CAGR between T1 and online survey and continuously measured variables of interest (assessed at T1).
